# Effects of pharmacological interventions on short- and long-term mortality in patients with takotsubo syndrome: a report from the SWEDEHEART registry

**DOI:** 10.1101/2023.08.31.23294927

**Authors:** P Petursson, E Oštarijaš, B Redfors, T Råmunddal, O Angerås, S Völz, A Rawshani, K Hambraeus, S Koul, J Alfredsson, H Hagström, H Loghman, R Hofmann, O Fröbert, T Jernberg, S James, D Erlinge, E Omerovic

**Affiliations:** Department of Cardiology, Sahlgrenska University Hospital, Gothenburg, Sweden; University of Pécs Medical School, Pécs, Hungary; Department of Cardiology, Falun Hospital, Falun, Sweden; Department of Cardiology, Skåne University Hospital, Lund, Sweden; Department of Cardiology, Linköping, University Hospital, Linköping, Sweden; Department of Cardiology, Umeå University Hospital, Umeå, Sweden; Department of Cardiology, Karolinska University Hospital, Stockholm, Sweden; Department of Cardiology, Södra Hospital, Stockholm, Sweden; Department of Cardiology, Örebro University Hospital, Örebro, Sweden; Department of Cardiology, Danderyd University Hospital, Stockholm, Sweden; Department of Cardiology, Uppsala University Hospital, Uppsala, Sweden

**Keywords:** takotsubo syndrome, takotsubo cardiomyopathy, stress cardiomyopathy, pharmacological interventions, prognosis, mortality

## Abstract

**Aim:** Takotsubo cardiomyopathy (TS) is a heart condition mimicking acute myocardial infarction. TS is characterized by a sudden weakening of the heart muscle, usually triggered by physical or emotional stress. In this study, we aimed to investigate the effect of pharmacological interventions on short– and long-term mortality in patients with TS.

**Methods:** We analyzed data from the SWEDEHEART (The Swedish Web-system for Enhancement and Development of Evidence-based care in Heart disease Evaluated According to Recommended Therapies) registry, which included 1,724 patients with TS among 228,263 unique individuals who underwent coronary angiography between 2009 and 2016. The majority of patients were female (77.0%), and the average age was 66 ± 14 years.

**Results:** Nearly half of the TS patients (49.4%) presented with non-ST-elevation acute coronary syndrome and a quarter (25.9%) presented as ST-elevation myocardial infarction. Most patients had non-obstructive coronary artery disease on angiography, while 11.7% had a single-vessel disease and 9.2% had multivessel disease. All patients received at least one pharmacological intervention, most used being beta-blockers or antiplatelet agents. All-cause mortality rates were 3.9% after 30-days and and 16.7% at long-term follow-up (median 877 days). Intravenous use of inotropes, diuretics, and orally administered digoxin were associated with increased mortality in TS, while ACE-I and statins were associated with decreased long-term mortality. Unfractionated and low-molecular-weight heparin (UH/LMWH) were associated with reduced 30-day mortality. However, medications such as ARBs, oral anticoagulants, P2Y12 antagonists, aspirin, and beta-blockers did not statistically correlate with mortality.

**Conclusion:** Our findings suggest that some medications commonly used to treat TS are associated with higher mortality while others with lower mortality. These results could inform clinical decision-making and improve patient outcomes in TS. Further research is warranted to validate these findings and to identify optimal pharmacological interventions for patients with TS.

## 1. Introduction

Takotsubo syndrome (TS) is a form of heart failure characterized by transitory left ventricular dysfunction and often resambles myocardial infarction (MI) in the acute phase ^1,2^. Several pathophysiological mechanisms, including excessive sympathetic stimulation and myocardial perfusion dysfunction, have been proposed to explain TS^1,3^. This syndrome often affects postmenopausal women and poses a differential diagnostic dilemma because it mimics MI. The diagnosis is based mainly on transient akinesia in apical segments detected by ventriculography or cardiac ultrasound in the absence of explanatory coronary angiography findings^1,3^.

Although the medical community first described TS in 1990^4^, its importance has significantly increased worldwide in the past decade. Since the cause of TS is still unknown, medical professionals have yet to establish the optimal treatment approach.

Extrapolating pharmacological treatments for acute MI and acute heart failure to TS can be dangerous and may result in severe consequences, including fatalities^5,6^. The optimal medical treatment for TS is still unknown, as no randomized controlled clinical trials have been conducted to evaluate the effects of pharmacological interventions.

Due to the transient nature of the disorder, it will be prudent to concentrate on supportive treatment^1,7^. However, pharmacological therapeutic approaches, such as beta-blockers, antiplatelets, statins, and angiotensin-converting enzyme inhibitors (ACE-I), are frequently used^8,9^. Medical professionals may assume that treatments used for other conditions, such as MI and acute heart failure will yield similar outcomes in TS^10^. However, the efficacy and safety of pharmacological treatments such as inotropes, beta-blockers, antiplatelets, statins, and ACE-I in TS remain unclear.

We sought to evaluate the effect of pharmacological treatment received during hospitalization and after discharge on short– and long-term survival in a large cohort of TS patients using comprehensive national Swedish registry data.

## 2. Methods

### 2.1. Study population

We included all hospitalized patients with TS who were reported to the the Swedish Coronary Angiography and Angioplasty Registry (SCAAR) between June 2009 and March 2016. TS was defined according to the position statement document from the TS task force of the European Society of Cardiology (ESC) Heart Failure Association^1^. All patients were included in the SCAAR and Swedish Register of Information and Knowledge about Swedish Heart Intensive Care Admissions (RIKS-HIA), which are part of the Swedish Web-System for Enhancement and Development of Evidence-Based Care in Heart Disease Evaluated According to Recommended Therapies (SWEDEHEART) registry. It provides a web-based platform dedicated to data collection from all angiographies and percutaneous coronary interventions (PCIs) performed in coronary catheterization laboratories in Sweden (n = 31) since 1989. The registry is financed by the county councils in Sweden and the Swedish state. To obtain information about the patients’ vital status, the database is continuously merged with the national population registry. The study was approved by the ethics committee at the University of Gothenburg (Diarienumber [Dnr]. 759-13, date of approval May 6, 2014).

### 2.2. Definitions

The SCAAR uses standardized definitions across all hospitals, and all information is entered directly into the database by interventional cardiologists. Since 2009, interventional cardiologists report TS based on the criteria (Table S1). The variables in the SCAAR and the RIKS-HIA registries are routinely validated against patient charts and considered highly accurate. Coronary artery disease is defined as the presence of stenosis >50% in any coronary artery.

*Endpoints:* We evaluated the association between the received pharmacological treatment and mortality at 30 days and after the long-term follow-up. The primary endpoint was mortality at 30 days. Vital status and date of death were obtained from the Swedish National Population Registry until March 7, 2016. The SCAAR was merged with the RIKS-HIA and the Swedish National Population Registry according to Swedish personal identification numbers. Because the use of unique identification numbers is mandatory, the population registry in Sweden has a high degree of completeness but is not reviewed or adjudicated to establish cardiac versus noncardiac causes of death.

### 2.3. Statistical analysis

Continuous variables are presented as a median and interquartile range, and categorical variables as frequencies. The normal distribution of variables was assessed by inspecting the distribution of values on histograms and by the Shapiro–Wilk test. Intergroup differences in continuous variables were tested by linear regression. Differences in categorical variables were tested by logistic regression.

*Imputation protocol:* Missing values in the dataset were imputed using the missRanger package in R, which utilizes random forest imputation to fill in missing values^11^. The missRanger method has been shown to produce accurate imputations while preserving the distribution of the data, making it a reliable approach in missing data imputation.

*Statistical modelling:* Cumulative mortality rates at 30 days and long-term are presented for patients within the entire cohort. After multiple imputation of missing data, multivariable Cox proportional hazards was used to adjust for differences in patient characteristics. All multivariable models were fit to the entire study cohort and were adjusted for variables in Table 1., Killip class, the use of intra-aortic balloon counter pulsation pump and duration of hospitalization. Treating hospital was included as a random effect variable to account for the clustering of patients within hospitals. To prevent immortal time bias when evaluation long-term mortality, the follow-up time began on the date after the discharge^12^. Every analysis used the commonly established criterion of statistical significance, which is a 2-tailed =.05. All statistical computations and data visualization were done in R, version 4.2.3 (R Foundation for Statistical Computing) and Stata, (release 18.0, College Station, TX: StataCorp LLC). Schoenfeld residuals were used to assess proportional hazards^13^. The variable hospital stay violated the proportional risks assumption. To address this violation, a time-varying Cox regression model^14^ was employed, allowing the effect of the variable to change over time. To account for nonlinear age-hazard relationships, age was represented as a spline with 3 knots.

**Table 1.**
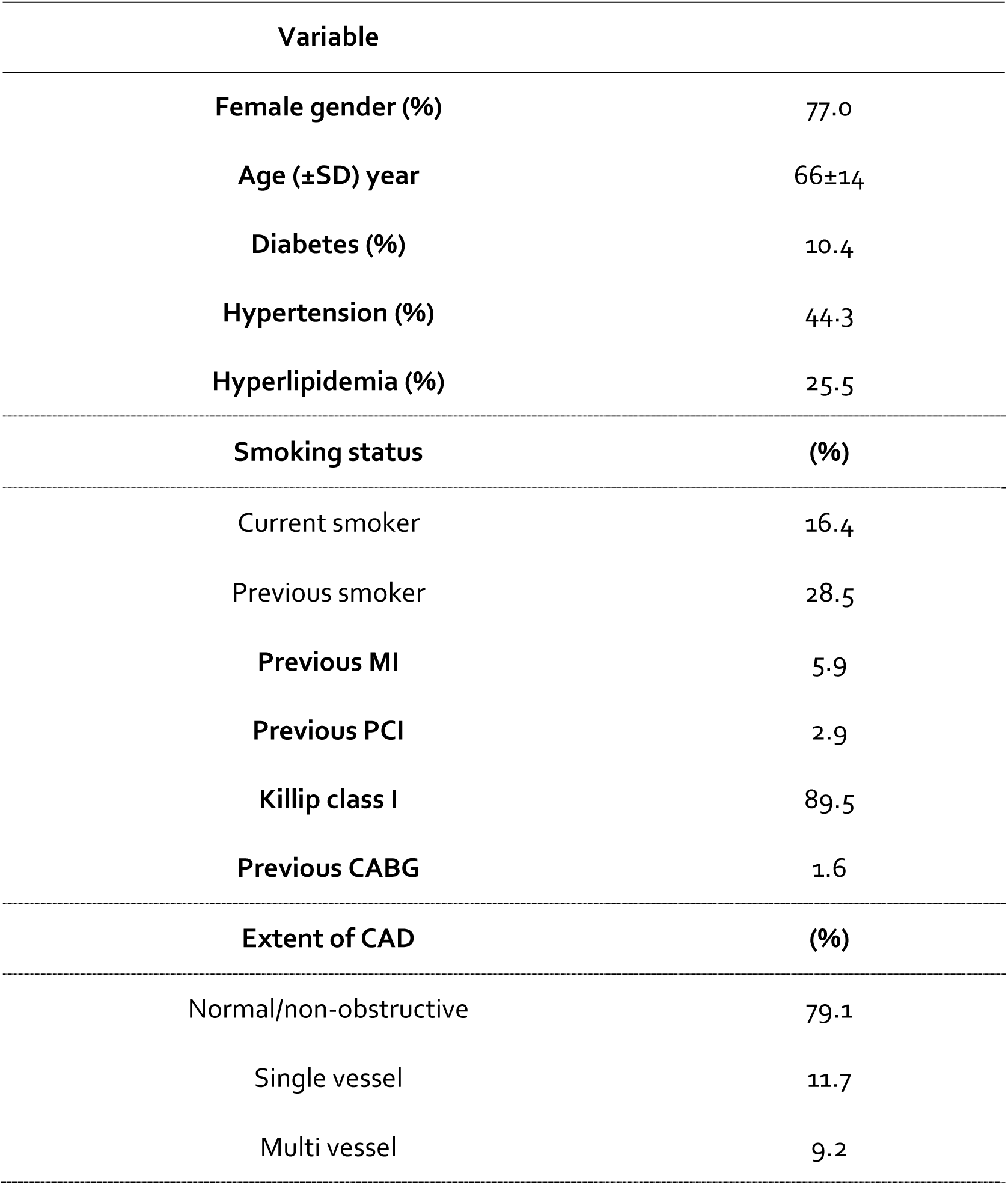
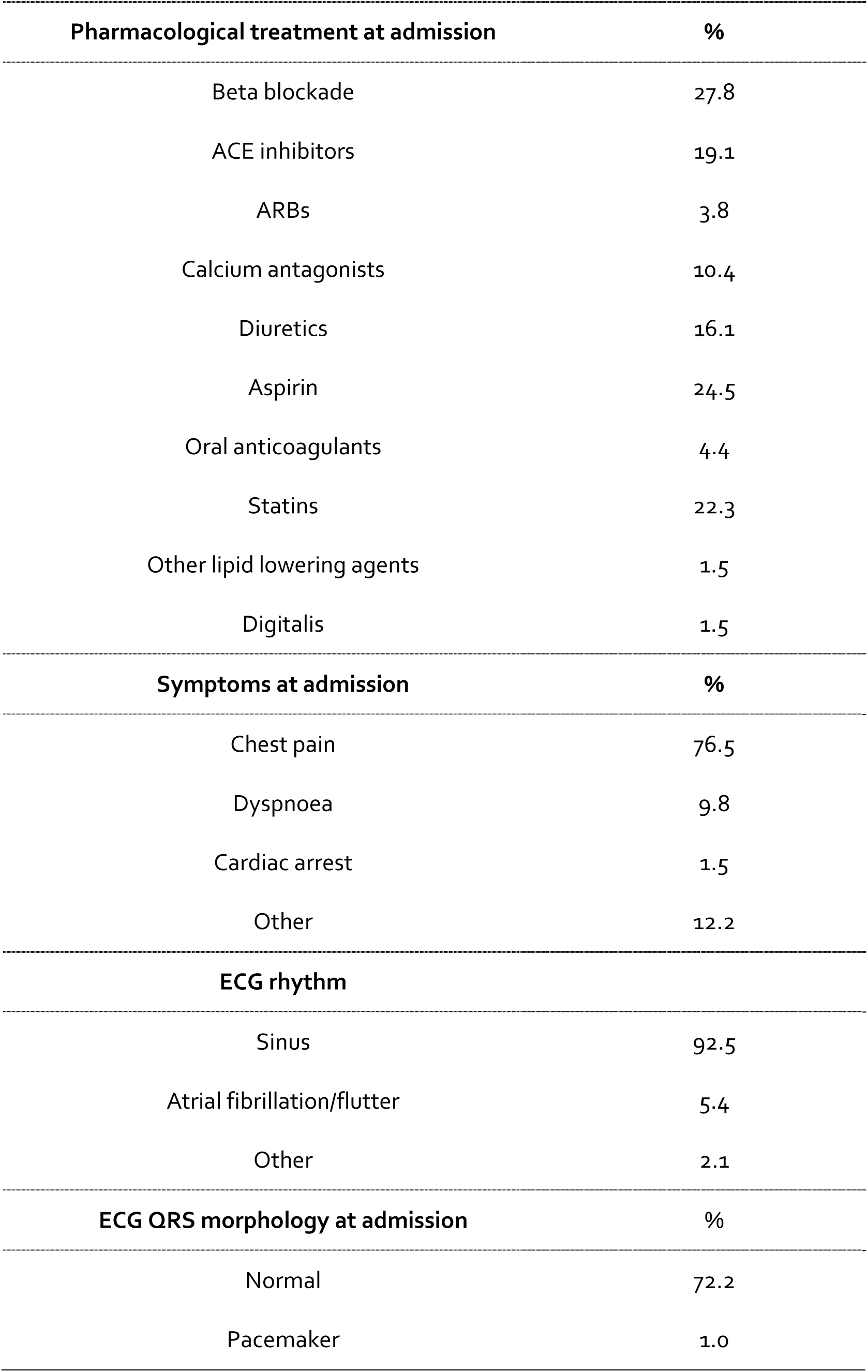
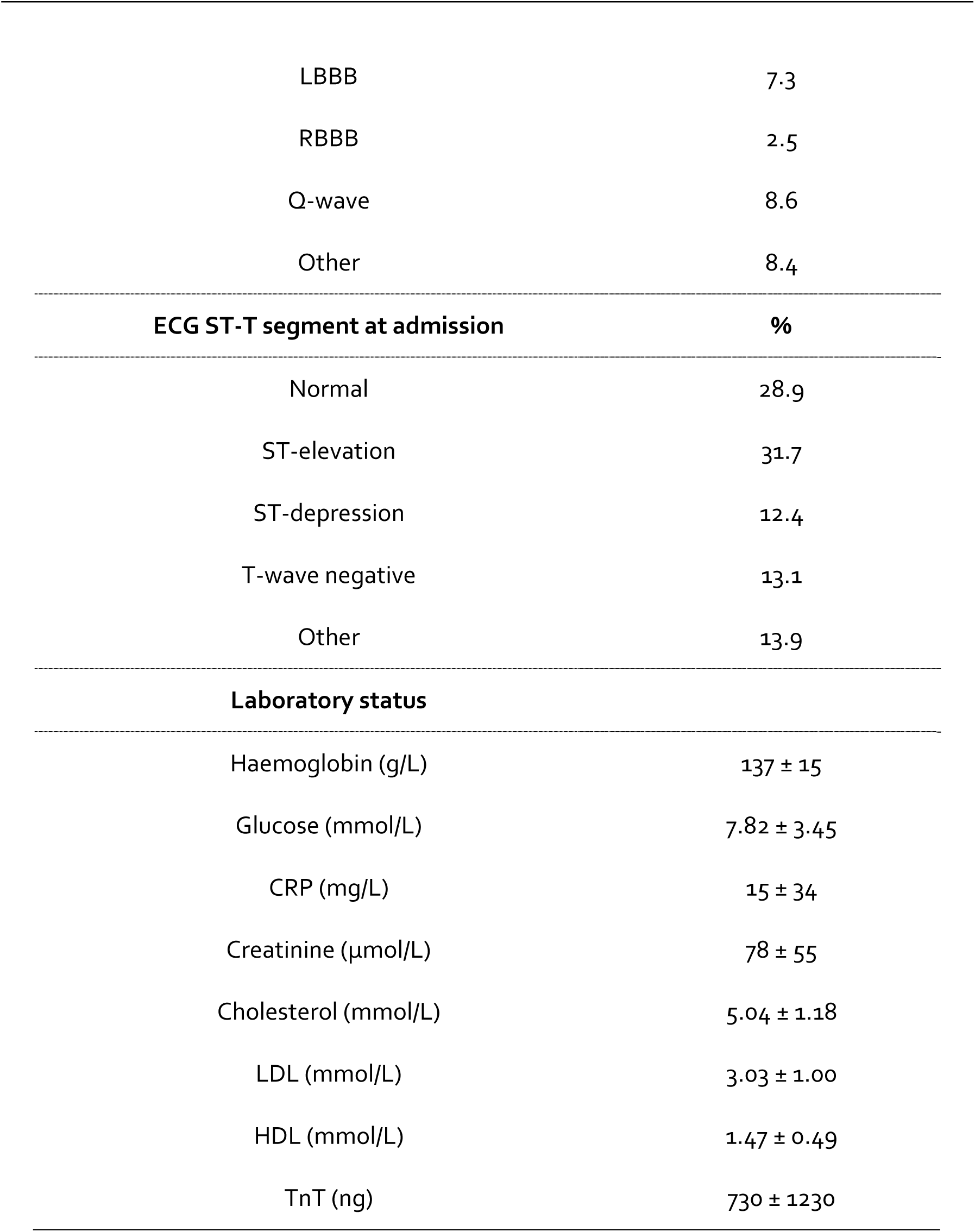
Patient’s characteristics.

*Sensitivity analyses and postestimation diagnostics:* Sensitivity tests were conducted to determine the extent to which an unmeasured variable may cause exposure to different treatments to lose statistical significance^15^. Goodness-of-fit for the models was assessed with the Harrell’s C test^16^. Multicollinearity between the variables in the model was evaluated by calculating the variance inflation factor (VIF).

## 3. Results

### 3.1. Patient characteristics and treatments

In total, we identified 1,724 patients with TS among 228,263 unique individuals who underwent coronary angiography and were reported to the SCAAR registry between 2009 and 2016 (Figure 1). The characteristics of the hospitalized patients, including their pharmacological treatment, are presented in Table 1. Seventy-seven percent of patients were female. The average age was 66±14 years, and diabetes (10.4%), hypertension (44.3%), and hyperlipidemia (25.5%) were the most frequent comorbidities. According to the smoking status, 16.4% were current smokers, while 28.5% were previous smokers.

**Figure 1.**
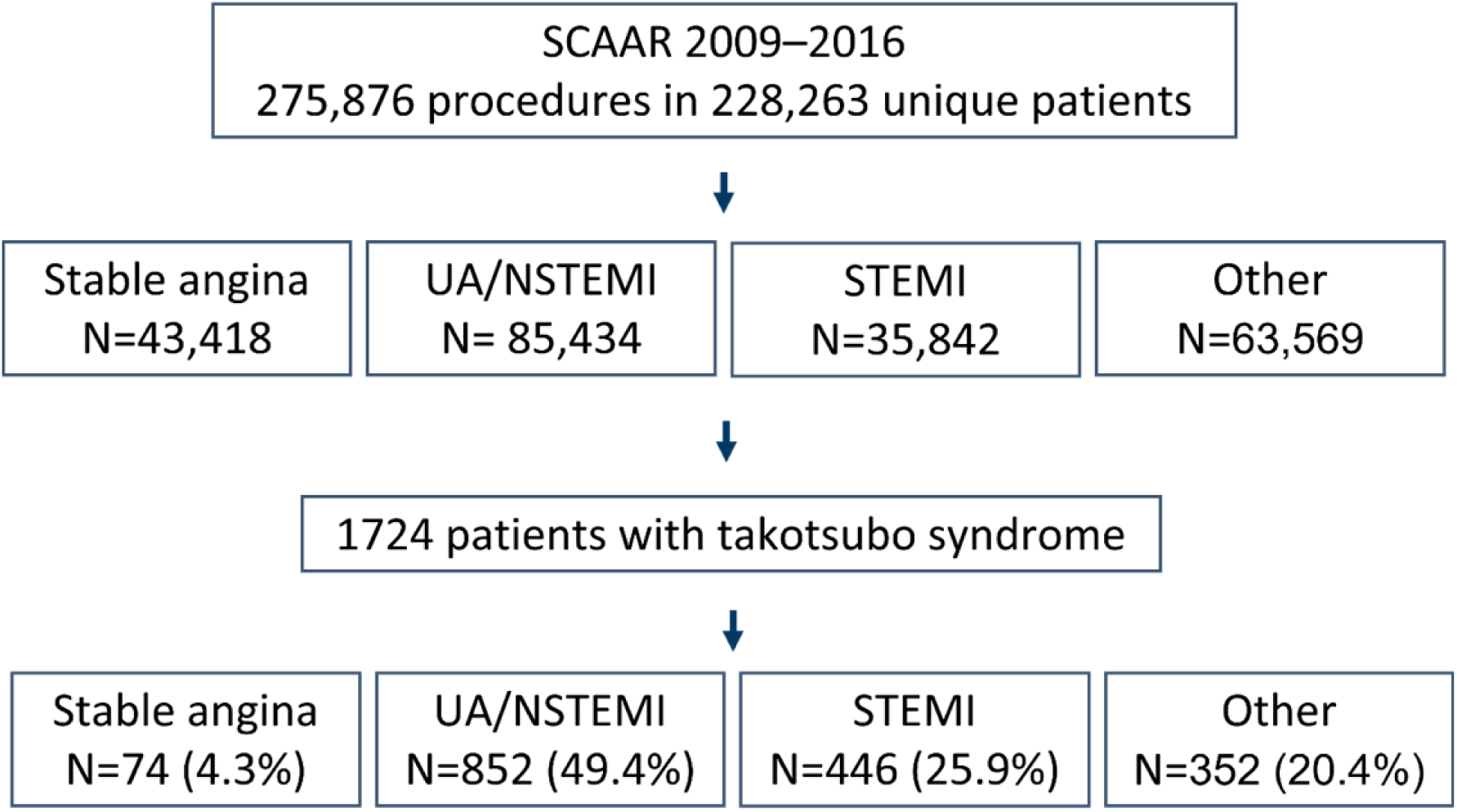
Patient selection chart. Patient selection chart for Takotsubo syndrome (TS) cases in a SCAAR. The study aimed to identify TS cases among patients who underwent coronary angiography and were reported to the SCAAR registry between 2009 and 2016. A total of 1724 patients with TS were identified among 228,263 unique individuals who underwent coronary angiography during the study period.

**Figure 2.**
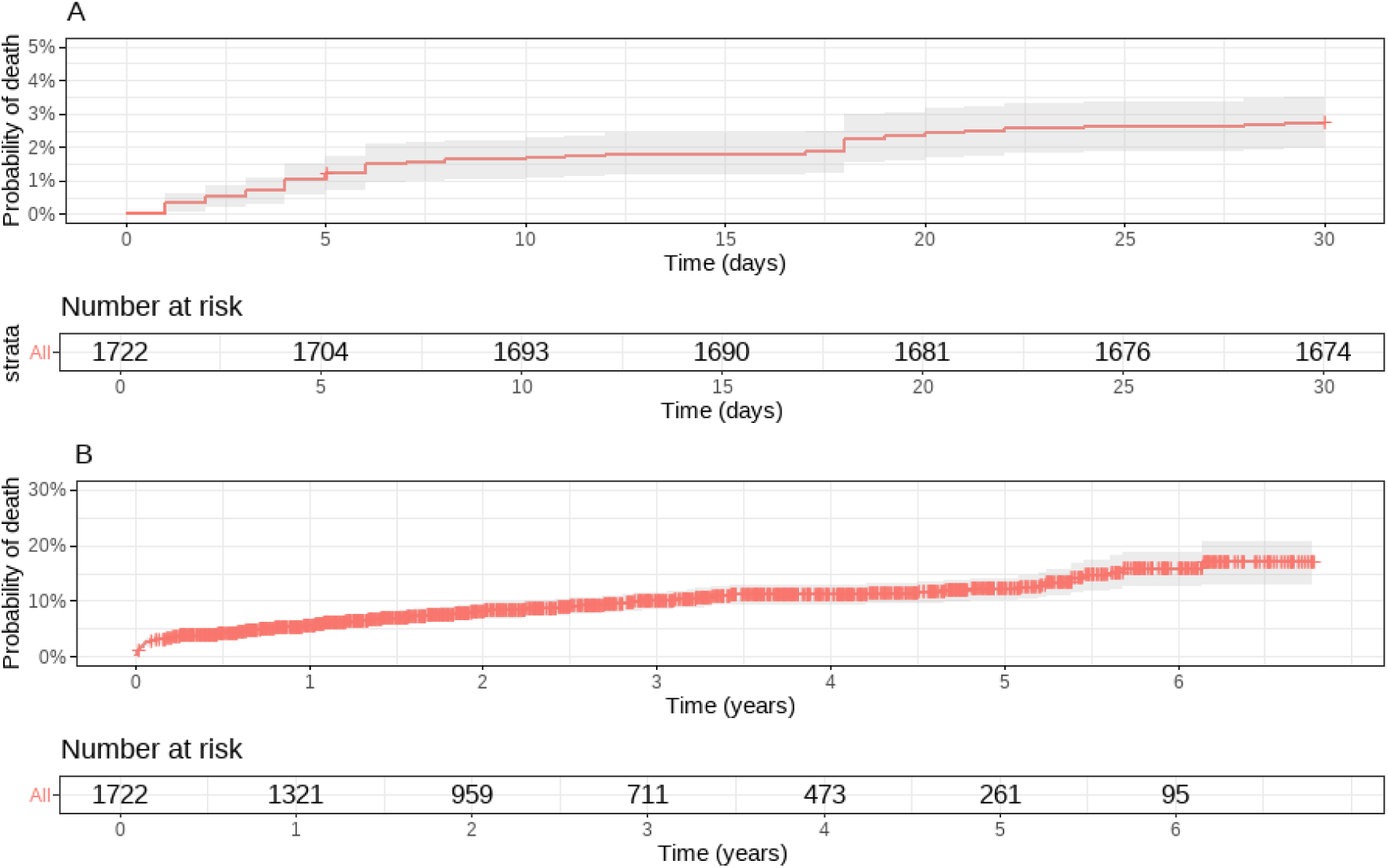
Kaplan-Meier curve for 30-day and long-term mortality in patients with TS. The Kaplan-Meier curve illustrating the 30-day (A) and long-term mortality (B) rates among patients diagnosed with Takotsubo syndrome (TS). The median follow-up duration of patients was 877 days (interquartile range [IQR]: 383-1544), and the crude all-cause mortality rates were found to be 3.9% and 16.7% at 30-day and long-term follow-up, respectively. Notably, the Kaplan-Meier curve demonstrated that the short-term and long-term mortality rates for TS patients are comparable to those observed among patients with myocardial infarction.

In half (49.4 percent) of the 1,724 patients the indication for coronary angiography was non-ST-elevation acute coronary syndrome, which includes unstable angina and non-ST-elevation MI. The remainder were reported as ST-elevation MI (25.9%), stable angina (4.3%), and other diagnoses (20.4%) including cardiac arrest, heart failure, valve disease, chest pain, arrhythmias. Killip I accounted for the majority of patients (88.9 %), followed by Killip II (7.8 %), Killip III (2.1 %), and Killip IV (1.2 %).

5.9 percent of patients had a history of MI, 2.9% had PCI, and 1.6% had undergone CABG treatment. Coronary angiography revealed that most patients (79.1 Only troponin levels exceeded the reference values among the results of the routine blood sample analysis (Table 1).%) had normal coronaries or non-obstructive disease, while 11.7% had a single-vessel disease and 9.9% had a multi-vessel disease.

Before hospitalization, a considerable number of patients were already using beta-blockers (27.8%), ACE inhibitors (19.1%), and calcium antagonists (10.4%). Additionally, 16.1% of patients were using diuretics, 24.5% were taking aspirin, and 4.4% were taking oral anticoagulants. In addition, 22.3% of patients were already taking statins, whereas a small percentage were getting other lipid-lowering medications and digoxin, respectively. Only troponin levels exceeded the reference values among the results of the routine blood sample analysis (Table 1).

All patients received at least one pharmaceutical intervention during hospitalization (Table 3). Most were given beta-blockers (77.8% orally and 8.3% intravenously) and antiplatelet medications (66.7% with aspirin and 43.6% with P2Y12 inhibitors). Other medications included unfractionated and low-molecular-weight heparin (UH/LMWH) (65.3%), oral anticoagulants (11.3%), ACE-I (55.5%), angiotensin receptor blockers (ARBs) (15.3%), statins (55.1%), diuretics (19.5% orally and 17.2% intravenously), inotropes (3.8%), and digoxin (1.3%). Intra-aortic balloon counterpulsation pump was used in 0.5%. The median duration of hospitalization was four days, with an interquartile range of 4.

**Table 2.** In-hospital treatment.

| Variable | % |
| --- | --- |
| Inotropes i.v. | 3.8 |
| Diuretics i.v. | 17.2 |
| Diuretics p.o. | 19.5 |
| Digoxin p.o. | 1.3 |
| UH/LMWH | 65.3 |
| Statins | 55.1 |
| ACE inhibitors | 55.5 |
| ARBs | 15.3 |
| Beta blockers i.v. | 8.3 |
| Beta blockers p.o. | 77.8 |
| Aspirin | 66.7 |
| Intra-aortic balloon counter pulsation pump | 0.5 |

**Table 3.** Results of sensitivity analyses assessing the impact of unmeasured confounding on treatment-mortality associations.

| Treatment | RR | P<sub>Treated</sub> (%) | P<sub>Untreated</sub> (%) |
| --- | --- | --- | --- |
| Inotropes i.v. | 10 | 70 | 10 |
| Diuretics i.v. | 2 | 80 | 10 |
| Diuretics p.o. | 2 | 30 | 20 |
| Digoxin | 2 | 60 | 10 |
| UH/LMWH | 2 | 50 | 90 |
| Statins | 2 | 20 | 30 |
| ACE inhibitors | 2 | 10 | 20 |

RR = relative risk for association between the unmeasured confounder and mortality.
**Interpretation:** When adjusting for unmeasured confounding, the upper or lower limit of the 95% CI for mortality would cross null losing significance with the specific treatment if the unmeasured confounder had a risk of mortality above the specific RR, considering the corresponding prevalence in the treated group (P<sub>Treated</sub>) and the untreated group (P<sub>Untreated</sub>). For example, to lose statistical significance as an independent predictor of death at 30 days, an unmeasured confounder would need to have a risk ratio above 10 with a prevalence of 70% in the inotrope-exposed group and 10% in the inotrope-unexposed group.

### 3.2. Clinical outcomes

The median follow-up time was 877 days (IQR 383–1544), with crude all-cause mortality rates of 3.9% after 30 days and 16.7% at long-term follow-up.

Pharmacological interventions associated with increased mortality in TS included intravenous use of inotropes and diuretics, and orally administered digoxin (Figure 3A). Treatment with statins and UH/LMWH was associated with lower 30-day mortality, while statins and ACE-I were associated with lower long-term mortality (Figure 3B). Medications that did not show a statistically significant association with mortality were ARBs, oral anticoagulants, P2Y_12_ antagonists, aspirin, and β-blockers (Figure 3C).

**Figure 3.**
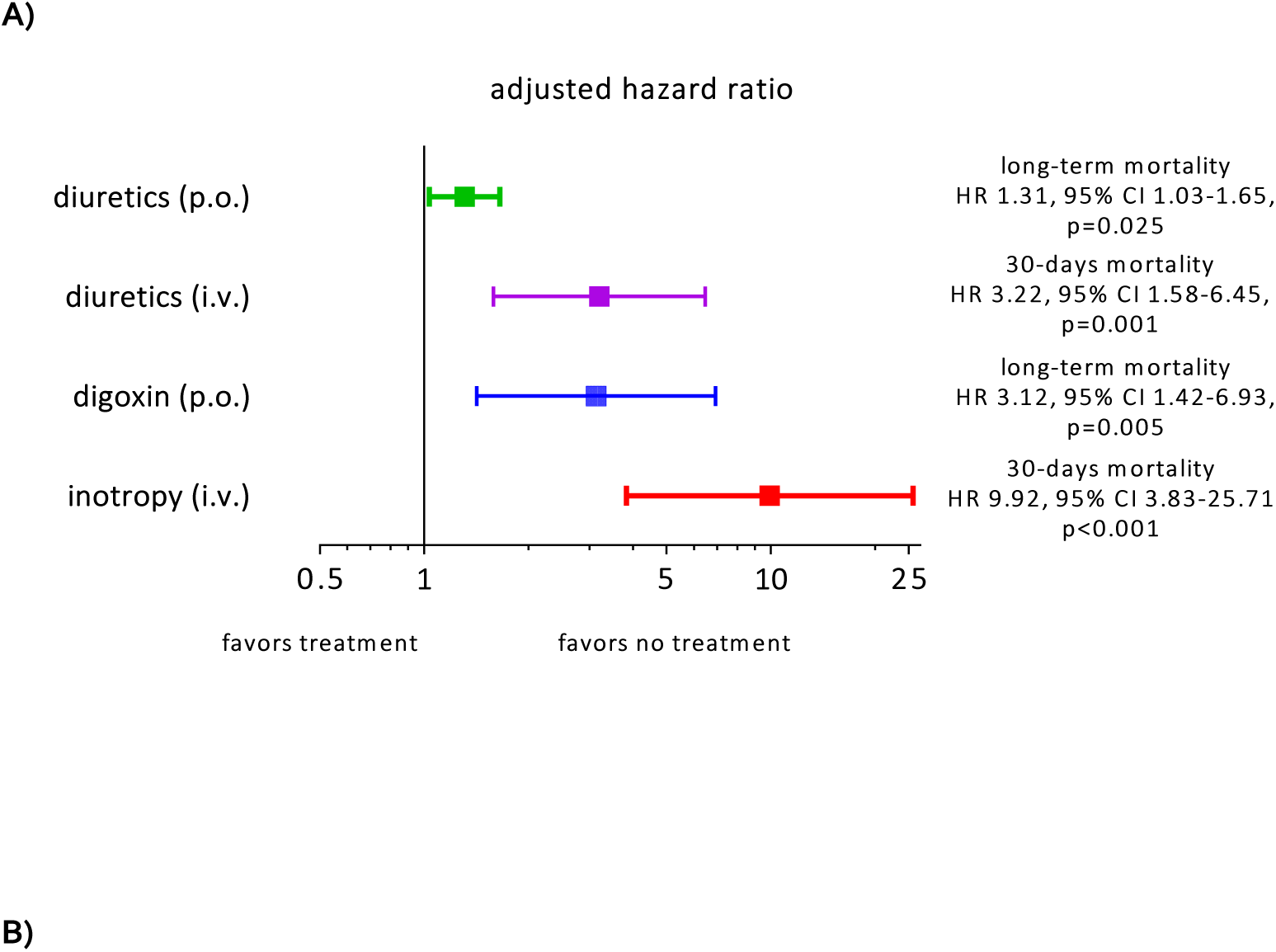
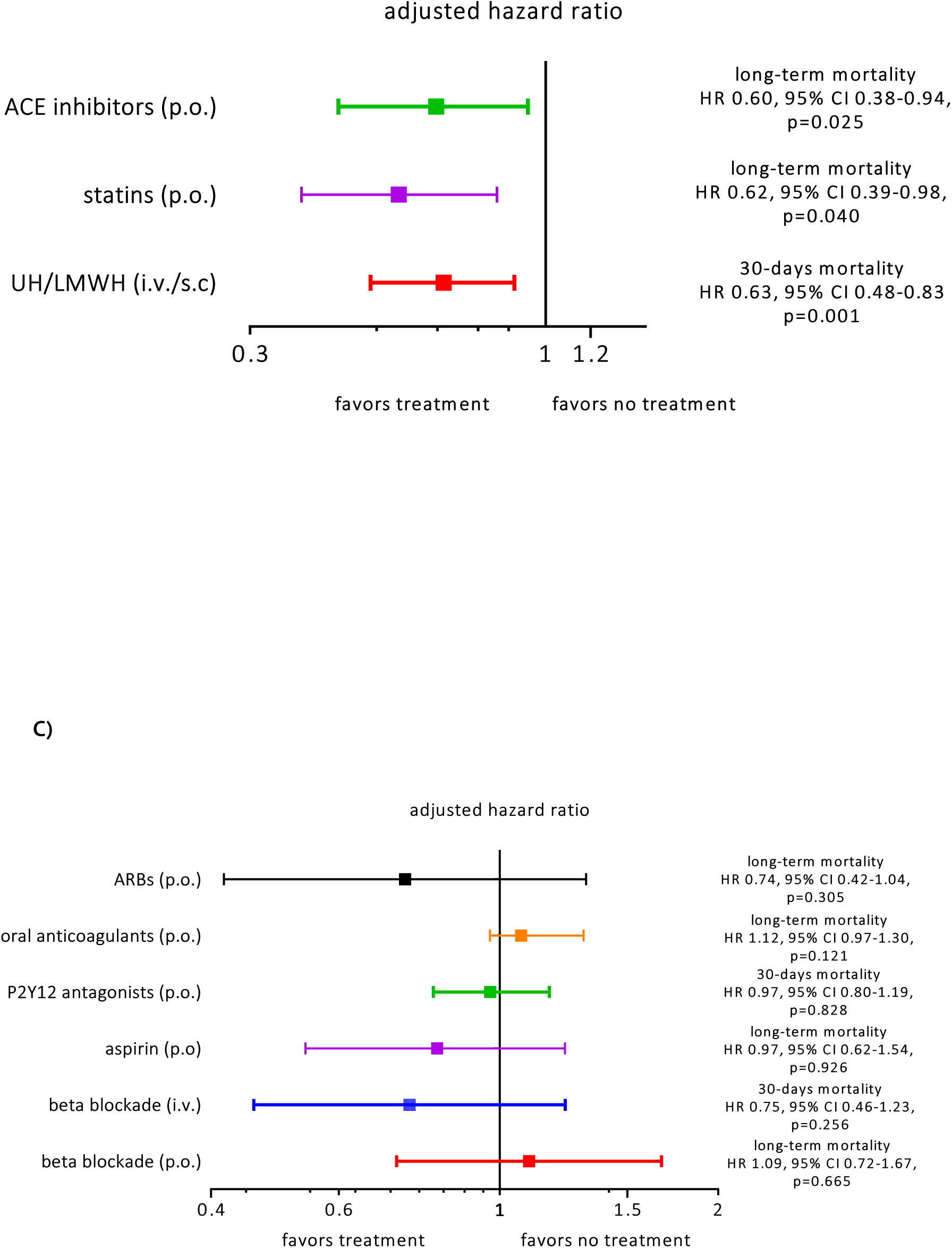
Pharmacological Interventions and Mortality Rates in Takotsubo Syndrome Patients: Associations and Outcomes. Figure 3 comprises of 3 panels displaying the associations between different pharmacological interventions and mortality rates among patients diagnosed with Takotsubo Syndrome (TS). Panel A indicates the use of intravenous inotrope, intravenous diuretics, and orally administered digoxin, which were associated with increased mortality rates. Panel B displays the use of ACE-I and statins, which were found to be associated with decreased long-term mortality rates. Furthermore, the use of UH/LMWH was associated with reduced 30-day mortality rates. Panel C shows that several medications, including ARBs, oral anticoagulants, P2Y12 antagonists, aspirin, and β-blockers (both intravenous and oral), did not exhibit a statistically significant association with mortality rates in TS patients. Due to wide 95% CI, risk estimates for ARBs, aspirin and orally and intravenously administered betablockade are inconclusive.

### 3.3. Sensitivity analyses and postestimation diagnostics

The time-varying Cox regression model showed good predictive accuracy with a Harrell’s C-index of 0.888, indicating the model’s good discrimination ability. We detected collinearity between the variable sex and age, with a VIF of 26 (a VIF > 5 indicating collinearity), and thus, we excluded sex from the model. There was no evidence of collinearity among the other predictor variables. The sensitivity analysis results are displayed in Table 3, indicating that the use of inotropes has the strongest association with mortality. To lose its statistical significance as an independent predictor of death at 30 days, an unmeasured confounder would need to have a risk ratio above 10. This is based on a prevalence of 70% in the inotrope-exposed group and 10% in the inotrope-unexposed group.

## 4. Discussion

Using data from the SWEDEHEART registry, we analyzed the effect of pharmacological therapy on mortality in a large cohort of 1,724 individuals with TS. Inotropes, diuretics, anticoagulants, ACE-I, and statins were significantly associated with short– and long-term mortality among TS patients. ACE-I, statins, and UH/LMWH were associated with lower mortality in TS, whereas the opposite was noted for inotropes, diuretics, and digoxin. Aspirin, beta-blockers, ARBs, P2Y12 antagonists, and oral anticoagulants were classified as neutral.

Previous studies^17–19^, have indicated that the mortality rates in patients with TS are comparable to those observed in patients with MI. However, there appears to be a significant disparity in mortality rates between different countries and even within the same country across different regions^19^. It is reasonable to hypothesize that differences in pharmacological interventions employed in managing TS may partly contribute to this variation ^19^.

Researchers have evaluated multiple therapies in various study designs, including retrospective cohorts, case series, and meta-analyses. Beta-blockers, ACE-I/ARBs, and antiplatelets are the most frequently evaluated drugs. Except for some reports showing possible therapeutic effect of levosimendan^20^, ACE-I/ARB^21,22^ and beta-blockers ^23^, the evidence for effective treatment of TS is scarce

One of the most important findings of our study is that inotropes are strongly associated with increased mortality in TS. It is reasonable to think that inotropes were mainly used in TS patients with hemodynamic instability. However, it is important to bear in mind that inotropes may aggravate heart failure in patients with TS. This is consistent with the prevailing theory concerning the pathophysiology of TS, where catecholamines play an essential role in forming typical akinesia in apical segments. There is solid evidence that administering catecholamines to humans^24^ and animals^25^ causes TS phenotype. Some data support that levosimendan, a phosphodiesterase inhibitor with both inotropic and vasodilatory effects, might benefit the recovery of TS^26^. However, caution should be taken with all interventions reducing afterload in TS because ∼25% of these patients may have left ventricular outflow tract obstruction^27^. Both inotropic and vasodilating treatment can be detrimental in patients with left ventricular outlflow tract obstruction and hemodynamic instability^28^. In addition, there is experimental evidence against using levosimendan in TS^29^. Our study confirms these observations, as utilizing inotropes was associated with the highest risk of dying within 30 days.

Increased risk of death was associated with the use of diuretics. In patients with acute heart failure, high-dose diuretics or aggressive diuresis may exacerbate renal function, electrolyte abnormalities, and central hemodynamics^30^. Diuretic-induced hypovolemia may be more harmful in TS patients with left ventricular outflow tract obstruction. Many patients with TS have also decreased peripheral vascular resistance^31^, and diuretic therapy may aggravate volume depletion and hemodynamic instability. Diuretics can also activate the sympathetic nervous system, which may be deleterious in TS as an overactivated sympathetic system, and high catecholamine levels are regarded to be central to TS pathophysiology^32^.

Antiplatelet therapy was assessed in a recently published meta-analysis^33^, with evidence suggesting its association with adverse outcomes; however, one study seemed to have influenced the conclusion. We found that P2Y12 inhibitor and aspirin were not associated with adverse outcomes, while anticoagulant therapy was associated with a beneficial outcome.

In our study ACE-I was associated with decreased long-term mortality while ARBs were not. A lack of power to show statistically significant association may explain this discrepancy as relatively few patients received ARBs. However, although commonly grouped because of their analogous inhibitory mechanism on the renin-angiotensin system (RAS), ACE-I and ARBs have different sites of action. ACEIs block the conversion of angiotensin I to angiotensin II by inhibiting the ACE enzyme, while ARBs block the binding of angiotensin II to the angiotensin II type 1 receptor. While both medications effectively reduce the activation of the RAAS system, ACEIs may also affect the breakdown of other substances, such as bradykinin, which can lead to an increase in vasodilation and may have additional cardioprotective effects, such as inhibition of the central sympathetic system. ARBs, conversely, are more selective in their target receptor and may have less effect on other pathways. Another explanation is related to differences in dosing and individual patient characteristics. ACEIs and ARBs may be prescribed at different doses depending on the patient’s characteristics, such as age, sex, comorbidities, and overall health status. These differences in dosing could contribute to differences in effectiveness and impact on survival. This observation is at least indirectly supported by meta-analysis in heart failure patients in which ACEIs, but not ARBs, reduced all-cause mortality and cardiovascular deaths^34^.

Despite being favorable in our assessment of TS, statins have not been well explored so far. To the best of our knowledge, no other investigation has reported their favorable connection with mortality in TS. Statins have anti-inflammatory properties, and TS is thought to be associated with the complex interplay of inflammatory processes. By reducing inflammation, statins may help moderate some underlying pathophysiological mechanisms involved in TS, improving outcomes. Secondly, statins are commonly prescribed for their cholesterol-lowering effects, but they may also have additional benefits on the cardiovascular system, such as improving endothelial function and reducing oxidative stress. These effects may help to improve overall cardiovascular health and reduce the risk of complications in patients with TS. Hypertension, diabetes, and hyperlipidemia are common comorbidities associated with TS. Statins are frequently prescribed for managing these comorbidities; therefore, patients with TS who are already taking statins may have better control of their underlying conditions, which may improve survival.

The data obtained from the SWEDEHEART registry is unique because of its nationwide coverage and high-quality input. Selection bias, usually attributed to observational studies, can be considered low in our study due to the inclusion of the entire population compared to sample-based studies. However, several limitations of the registry need to be addressed. Because of the registry’s observational and non-experimental nature, no definitive causal relationship between pharmacological interventions and mortality can be made. The lack of detailed information on the route of administration, daily doses, and length of treatment makes interpretations of any association between a drug class and outcome challenging. To increase the level of evidence and establish a causal relationship between the pharmacological interventions and mortality in TS, more research should be conducted, ideally with patients randomly assigned to a pre-specified intervention in a randomized controlled trial (RCT). The currently ongoing BROKEN SWEDEHEART study^35^ with 2 x 2 factorial design will evaluate whether adenosine/dipyridamole and apixaban improve contractile recovery and decrease major adverse cardiovascular events at 30 days.

Even though TS is self-healing in most cases, it can manifest as life-threatening heart failure in the worst-case scenario. Some of the results of our analysis are consistent with previously reported observations, supporting the hypothesis that specific pharmacological treatments may be beneficial while others may be harmful. To clarify this matter, it is necessary to conduct high-quality RCTs and observational studies. In the meantime, it is advisable to exercise caution when implementing pharmacological interventions, adhering to the fundamental principle of “primum nil nocere” (first, do no harm).

## Data Availability

Data will not be available to public due to the restriction stipulated by Swedish law.

